# Covid-19 Vaccine Efficacy: Accuracy, Uncertainty and Projection of Cases

**DOI:** 10.1101/2020.12.16.20248359

**Authors:** Wenjiang Fu, Jieni Li, Paul Scheet

## Abstract

**Background:** Two vaccine candidates for coronavirus disease 2019 (Covid-19) have been announced by Pfizer-BioNTech and Moderna with above 90% efficacy. The efficacy of each vaccine changes between reports with no accuracy assessment.

**Methods:** We examined data in both vaccine trials, provided 95% confidence intervals, and projected the cases that would be prevented in communities of multi-million population.

**Results:** The 95% confidence intervals reveal that the true vaccine efficacy could be as low as 86% for stated efficacy of 94.4% in an interim report, indicating the inaccuracy and uncertainty of efficacy point estimate. Both vaccines achieve an efficacy above 89% by the 95% confidence interval in updated reports. The Moderna vaccine would prevent more than 50,260 cases in communities of 1 million people with 1 year exposure.

**Conclusions:** Point estimates of vaccine efficacy transmit limited information. Corresponding statements of uncertainty, such as confidence intervals, should be provided and included in discussions of societal impact. The Covid-19 vaccines announced to date would prevent a substantial number of cases even at lower ends of the intervals.

## 1. Introduction

Vaccination is crucial in blocking fast spread of deadly infectious diseases, such as the highly contagious Covid-19, especially when effective treatment or cure of the disease does not exist. Efficacy is one of the key indices to evaluate vaccine. It measures the effect of vaccination by calculating the percentage reduction of risk among the vaccinated subjects out of the unvaccinated in a double-blind placebo-controlled randomized clinical trial [1, 2, 3].

In the development of vaccines for Covid-19, Pfizer and BioNTech first announced their interim results with an efficacy above 90% on November 9, 2020 [4], and later updated the efficacy to 95% on November 18, 2020 [5]. Meanwhile, Moderna claimed a vaccine efficacy of 94.5% on November 16, 2020 [6], and updated the data on November 30, 2020 claiming 100% efficacy in preventing severe Covid-19 cases [7].

How accurate are these vaccine efficacies? Why does the efficacy fluctuate? What do deviations from these point estimates of vaccine efficacies imply for general population? In this paper, we provide explanations, quantify the uncertainty of vaccine efficacy using confidence interval (CI), a statistical measure of the accuracy, and further project the number of cases a vaccine would prevent in a community of multi-million people based on the current clinical trial data.

## 2. Randomness of Vaccine Efficacy and Confidence Interval

In examining vaccine efficacy, a doubled-blind placebo-controlled clinical trial is often conducted, as in the Pfizer-BioNTech and the Moderna studies. In such trials, a moderate or large number of participants are recruited over a period of months and randomly assigned to one of two trial groups (or two arms) to ensure the comparability of subjects between the two groups. One group is called treatment group and its subjects receive the study vaccine. The other group is called control group and its subjects receive placebo with no generic study medication, mimicking the population in the communities who would not receive vaccine treatment. The efficacy is then calculated to be (1 − *RR*), with *RR* being the risk ratio, a ratio of the disease rates of the vaccine group *R*_*v*_ to the control group *R*_*u*_. Hence, the smaller the rate among the vaccinated relative to the unvaccinated (*R*_*v*_/*R*_*u*_), the higher the vaccine efficacy (1 − *RR*) [2].

Since the participants are recruited over time to the clinical trial, the disease risk of each group is calculated with the number of cases diagnosed divided by total exposure of the group, and the total exposure is the sum of the individual exposure time. For example, among 100 participants of the vaccine group, if 20 have received the treatment for 5 months, 20 for 4 months, 20 for 3 months, 25 for 2 months, and 15 for 1 month, the total exposure is calculated as 20 × 5 + 20 × 4 +20 × 3 + 25 × 2 + 15 × 1 = 305 person-month, or 25.417 person-year. If 3 subjects among the 100 are diagnosed at the end of their exposure, the risk is 3/25.417 = 0.118, or 118 per 1000 person-year.

Although the total exposure of each group increases over time, it is not random because it is designed to increase over time by the study investigator. However, the number of cases diagnosed in each group is random, which is subject to the chance of the participants being exposed to the disease in the environment, individual immune response to the virus and the vaccine, which may depend on a participant’s age, race, and sex. Hence the risk of each group is random and fluctuates over time, and so does the vaccine efficacy estimate.

Such efficacy calculated based on clinical trial data represents a realization of the true efficacy among the participants and can be thought of to follow a probability distribution. It can also be used to estimate of the true efficacy. In assessing the accuracy of such an estimate, a CI can be constructed based on the trial data. For this purpose, we calculate the variance of the logarithm of the risk ratio log(*RR*), and construct a 95% CI based on a Poisson distribution assumption. It assumes that the number of cases *X* in the vaccine group of exposure *N*_*v*_ follows a Poisson distribution. Similarly, the number of cases *Y* in the placebo group of exposure *N*_*u*_ also follows a Poisson distribution with a different mean. The risk ratio is calculated as *RR* = (*X*/*N*_*v*_)/(*Y*/*N*_*u*_), and a 95% confidence interval of the efficacy is

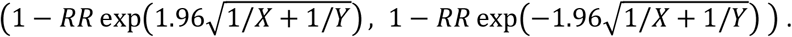

Details of the CI procedure and its derivation are provided in the Methods section.

We apply the above CI to the vaccine trial data. Although the risk may potentially depend on the age, race, and sex of the participants in practice, we assume the risk is homogeneous for participants in each group because no detailed demographic information has been released by either study of Pfizer and BioNTech, or Moderna. Once such information becomes available, it is straightforward to adjust the efficacy and CI with stratified risk ratio method.

We examine the vaccine efficacy in Table 1 assuming identical exposure between the two groups. Based on Pfizer and BioNTech release on November 30, 2020, the results of *X* = 8 and *Y* = 162 cases in the vaccine and control groups, respectively, achieve an efficacy of 95.06% with a 95% CI (89.96, 97.57). The interpretation of this CI is that there is a 95% confidence that the true vaccine efficacy is between 89.96% and 97.57%. Likewise, the results of Moderna’s interim results with *X* = 5 and *Y* = 90 on November 16, 2020 achieve an efficacy of 94.44% with a 95% CI is (86.33, 97.74). The update with *X* = 11 and *Y* = 185 on November 30, 2020 achieves an efficacy of 94.05% with a 95% CI (89.08, 96.76). Hence the slight drop of Moderna’s vaccine efficacy is neither statistically significant nor substantial.

**Table 1.** Efficacy and 95% Confidence Interval based on Press Release Data*

| Company | Release date | Cases ($X, Y$) | Participants | Efficacy | 95% CI |
| --- | --- | --- | --- | --- | --- |
| Pfizer - BioNTech | Nov 18, 2020 | (8, 162) | 43000 | 95.06% | (89.96, 97.57) |
| Moderna | Nov 16, 2020 | (5, 90) | 30,000 | 94.44% | (86.33, 97.74) |
| Moderna | Nov 30, 2020 | (11, 185) | 30,000 | 94.05% | (89.08, 96.76) |

\*Efficacy is calculated assuming identical exposure in vaccine and control groups.

To understand how the exposure ratio between the vaccine and control groups *N*_*v*_/*N*_*u*_ influences the efficacy, we calculate the efficacy and the 95% CI with fixed numbers of cases *X* and *Y* but varying ratio *N*_*v*_/*N*_*u*_, and find that they increase slightly with the ratio *N*_*v*_/*N*_*u*_ but remain stable within a reasonable range [0.9, 1.1] of deviation from identical exposure. See supplementary material for more details. Hence it is reasonable to assume identical exposure between the two groups of clinical trials.

## 3. Projection of Cases Prevented by the Vaccine

To understand how the vaccines will help prevent the disease spread in the communities, we project the number of cases that would be prevented in a community of multi-million people. For given intensities of the disease among the vaccinated and unvaccinated groups, the difference *λ*_*u*_ − *λ*_*v*_ is the intensity of cases the vaccine would prevent. Hence the number of cases to be prevented follows a Poisson distribution with a mean of the above intensity difference by the exposure *Pois*((*λ*_*u*_ − *λ*_*v*_)*N*). For the purpose of illustration, we project the number of cases that would be prevented by the Moderna vaccine among a community of multi-million people, and calculate the middle 95% range of the number of cases. Similar projection can be done for the Pfizer-BioNTech vaccine.

Figure 1 shows the middle 95% range of the projected number of cases for the exposure *N* of 1, 2, 4, 6, 8 and 10 million person-year. It is shown that the number of cases that would be prevented is proportional to the population exposure with a slope of about 5,026.7 cases per 100,000 person-year. For example, among 1 million people with 1 year exposure, the vaccine would prevent 50267 cases. Detailed calculation of the risk intensity is provided in the supplementary material.

**Figure 1.**
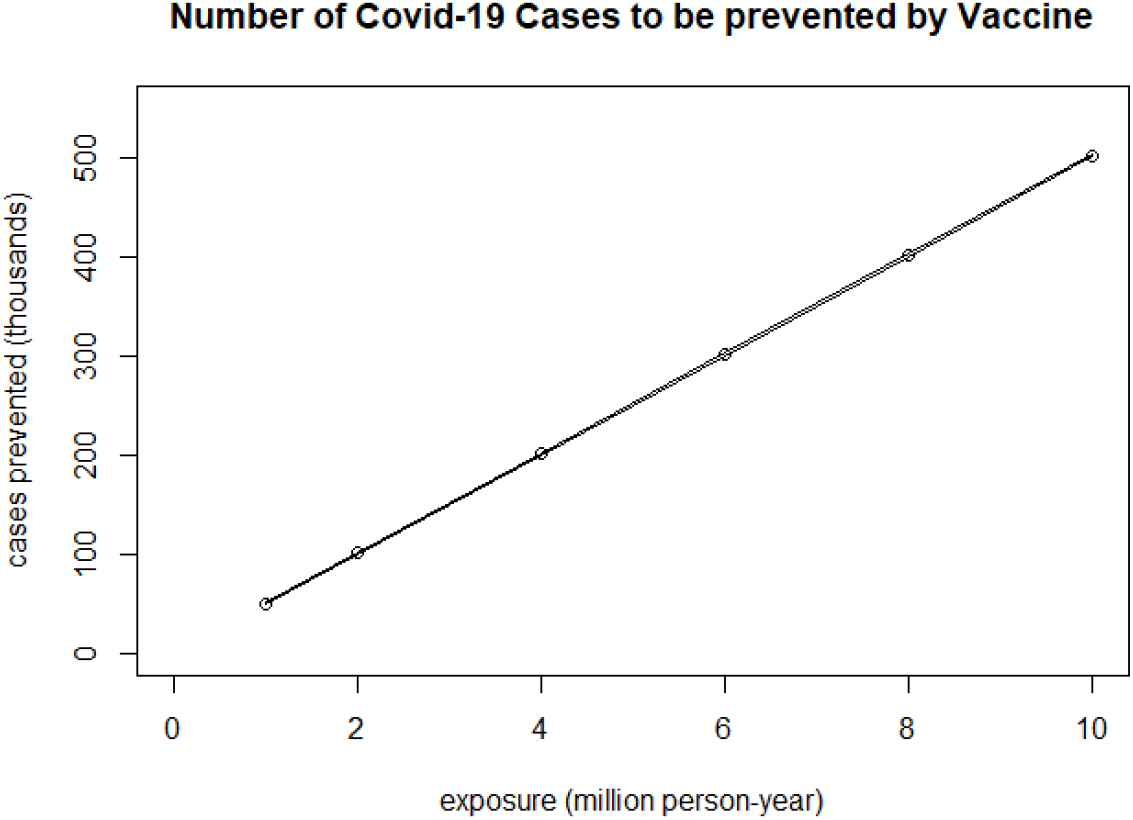
Projected number of Covid-19 cases that would be prevented by the Moderna vaccine.

## 4. Methods

### 4.1 Confidence interval

In the literature, numerous methods of constructing confidence intervals of risk ratio have been studied based on binomial distribution. But their pros and cons have made no consensus yet [9,10,11]. Here we show that the binomial distribution is less appropriate, whereas a Poisson distribution allows for a better reflection of reality. Since the participants are recruited over time, they have different exposures by the time the vaccine efficacy is calculated. Hence the individual probability of developing the disease varies within a group, which violates the equal probability assumption of the binomial distribution. A Poisson distribution is well known to approximate binomial distribution in large trials, and its assumption on rare event is valid given the small percentages of cases in both trials, 8/21500 and 162/21500 of the Pfizer-BioNTech vaccine and 11/15000 and 185/15000 of the Moderna vaccine.

Assume the number of cases *X* follows a Poisson distribution *Pois*(*λ*_*v*_*N*_*v*_) with mean being the intensity of the risk among the vaccinated *λ*_*v*_ by the total exposure *N*_*v*_. Similarly, *Y* follows a Poisson distribution *Pois*(*λ*_*u*_*N*_*u*_) with mean being the intensity of risk *λ*_*u*_ by the exposure *N*_*u*_ among the unvaccinated. For the trial data, the rates are *X*/*N*_*v*_ and *Y*/*N*_*u*_ for the vaccinated and unvaccinated, respectively. The risk ratio is calculated as *RR* = (*X*/*N*_*v*_)/(*Y*/*N*_*u*_). We derive the variance estimate of log(*RR*) for statistical convenience using the Delta method as follows [12].

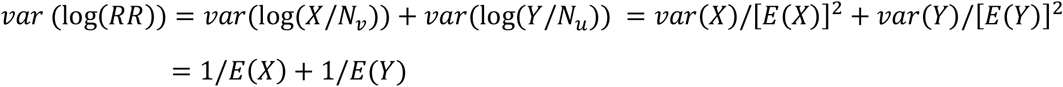

where the expected values *E*(*X*) and *E*(*Y*) can be estimated with *X* and *Y*. Hence a 95% CI of log(*RR*) is 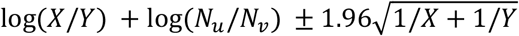, and a 95% CI of the vaccine efficacy is thus

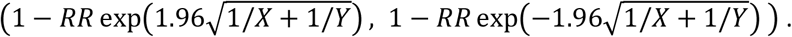

To validate the 95% CI through Monte Carlo simulations, the numbers of cases out of the total specified exposure of each of the two groups are generated from Poisson distributions with specified intensities and exposures, the efficacy and CI are calculated with the generated numbers of cases in the vaccine and control groups. The procedure is repeated *m* times (*m* = 1000, 5000 and 8000). The true efficacy is calculated with the specified intensity of each group as the risk. The proportion of the number of CIs out of *m* containing the true efficacy is examined. Proportions close to 95% confirm the validity of the CI.

### 4.2. Calculation of risk intensities for projection

The risk intensities are calculated based on the numbers of cases and exposure in the Moderna vaccine study. The Moderna trial started on July 27, 2020 and completed the recruitment of 30,000 participants about 12 weeks later by October 22, 2020 [6, 12]. Assuming half of participants are assigned to each group and the participants have been evenly recruited over 12 weeks. The overall exposure in each group during the 18 weeks (up to November 30, 2020) is calculated as 15000 × (12/2 + 6)/52 = 3461.54 person-year. The intensity of risk that the vaccine would prevent is (185 − 11)/3461.54 = 0.050267. This intensity multiplied by the exposure is the mean number of cases prevented.

## SUPPLEMENTARY MATERIAL

### S1. Validity of 95% CI

Table S1 displays the proportions of CIs that contain the true efficacy calculated from the known risks in the Monte Carlo simulations based on the number of cases in the Pfizer-BioNTech and Moderna reports. The validity of the 95% CIs is confirmed by the proportions close to 95%.

**Table S1.** Proportion of 95% CI containing true vaccine efficacy value in simulations*

| Simulation runs | 1000 | 5000 | 8000 |
| --- | --- | --- | --- |
| Pfizer - BioNTech | 0.9540 | 0.9560 | 0.9473 |
| Moderna | 0.9620 | 0.9516 | 0.9509 |

\*True efficacy is calculated based on known risk intensities that generate data for the vaccine and control groups.

### S2. Ratio of population exposure N_v_/N_u_ on the efficacy and 95% confidence interval

The variations of the efficacy and 95% confidence interval are examined and illustrated in Figure S1 for both vaccines. It is concluded that the vaccine efficacies and confidence intervals remain stable within a reasonable range [0.9, 1.1] of the deviation from identical exposure in the two clinical trial groups.

**Figure S1.**
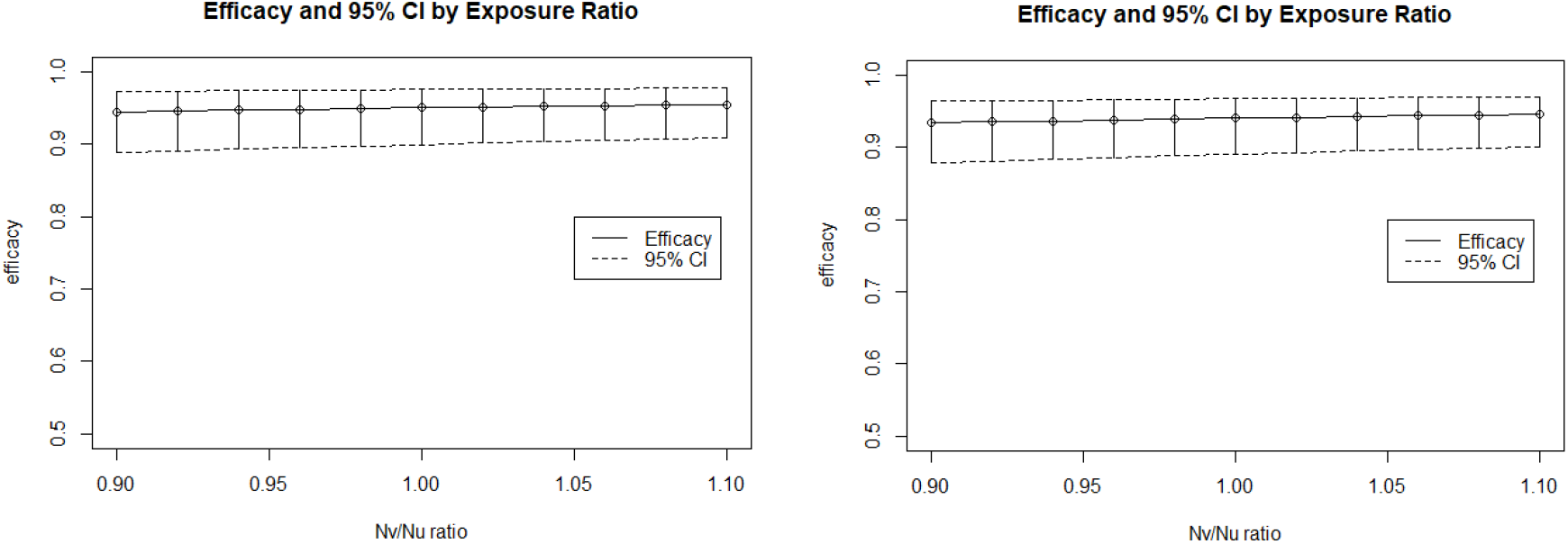
Efficacy and 95% confidence interval by varying exposure ratio. Left panel: Pfizer-BioNTech vaccine; Right panel: Moderna vaccine

## Data Availability

Publicly available.

